# Prevalence and pre-disposing factors of *helicobacter pylori* among patients with gastro-intestinal symptoms attending Mulago Hospital, Kampala, Uganda

**DOI:** 10.64898/2026.02.23.26346905

**Authors:** Racheal Twikirize, Phillip Wanduru, Tumwine Gabriel, David Musoke

## Abstract

**Background:** Comprehensive data on the prevalence of *Helicobacter pylori* infection and its associated risk factors among patients with gastrointestinal symptoms remain limited. Generating this evidence would help inform clinical management and improve antibiotic stewardship. *H. pylori* infection affects a substantial proportion of the global population, with prevalence varying widely across regions. In Uganda, previous studies have documented the presence of *H. pylori* infection. However, data specific to symptomatic patients are scarce. This study therefore aimed to determine the prevalence of *H. pylori* infection and associated factors among patients with gastrointestinal symptoms attending Mulago National Referral Hospital in Kampala, Uganda.

**Methods:** A cross-sectional study was conducted among 353 patients with gastrointestinal symptoms attending Mulago Hospital. Data on socio-demographic characteristics, lifestyle and dietary habits, and medical history were collected using a semi-structured questionnaire. *H. pylori* infection status was determined using stool antigen tests. Proportions were used to determine the prevalence of *H. pylori,* and associated factors analyzed using STATA version 14 software by performing bivariate and multivariate analyses.

**Results:** Among the 353 participants, majority were between 16 and 25 years old (69%), female (58%), and residing in peri-urban areas (74%). The prevalence of *H. pylori* infection in this population was 308 (87.3%). Multivariate analysis showed that *H. pylori* infection was significantly associated with having more than five income dependents (aPRR = 1.104, 95% CI: 1.025–1.189, p = 0.008), a history of previous *H. pylori* treatment (aPRR = 3.459, 95% CI: 2.138–5.595, p < 0.001), and a family history of *H. pylori* infection or gastrointestinal ulcers (aPRR = 1.135, 95% CI: 1.055–1.221, p = 0.001).

**Conclusion:** This study demonstrated a high prevalence of *Helicobacter pylori* infection among patients presenting with gastrointestinal symptoms, with nearly nine out of ten individuals testing positive. The high burden observed suggests that routine screening for *H. pylori*, or carefully guided empirical treatment, may be clinically justified in symptomatic patients. These findings underscore the need for integrated clinical and public health strategies to improve diagnosis, treatment, and prevention of *H. pylori* infection in this setting.

## Introduction

*Helicobacter pylori* (*H. pylori*) is a motile, gram-negative, urease-producing bacterium that colonizes the gastric mucosa, most often acquired during childhood (1). Globally, more than half of the population is infected, making it one of the most common chronic bacterial infections (1, 2). Its public health relevance stems from its established association with chronic gastritis, peptic ulcer disease, gastric adenocarcinoma, and mucosa-associated lymphoid tissue (MALT) lymphoma, leading the International Agency for Research on Cancer to classify it as a Class I carcinogen. (3, 4) Although infection can persist for life if untreated, only a minority of individuals develop clinically significant disease, while the majority remain asymptomatic (5). The diversity of bacterial strains and their virulence factors—such as CagA and VacA—plays an important role in determining disease outcomes. (6)

In many regions with high *H. pylori* prevalence, including East Asia, West Asia, and Africa, infection patterns exhibit notable epidemiological variations (1, 2). One widely discussed phenomenon is the “African and Asian enigma,” in which populations with high infection rates—particularly in Africa—show relatively low incidences of gastric cancer and peptic ulcer disease. (6, 7) Several hypotheses have been proposed to explain this disparity, including differences in dominant *H. pylori* strains, host genetic susceptibility, coexisting parasitic infections, nutritional status, and environmental exposures. (6-8) In sub-Saharan Africa, socioeconomic factors such as poor sanitation, household overcrowding, limited access to clean water, and high population density further contribute to transmission, likely through oral–oral, gastro–oral, or fecal–oral routes. (9) Despite these challenges, local epidemiological patterns remain incompletely understood.

In Uganda, *H. pylori* infection is recognized as a significant clinical concern, yet comprehensive data on its determinants remain limited. Although diagnostic methods such as stool antigen testing, urea breath tests, and serology are available, many health facilities rely on serology, which may lead to misclassification due to persistent antibodies. (10) Existing studies have documented high infection rates, but evidence on context-specific risk factors—such as household characteristics, prior treatment history, environmental exposures, and family-level clustering—remains scarce (7,11). This lack of detailed epidemiological data restricts effective prevention and management strategies. Therefore, this study aimed to determine the prevalence of *H. pylori* infection and identify associated risk factors among patients with gastro intestinal symptoms attending Mulago hospital in Kampala, Uganda.

## Methods and materials

### Study setting, population and design

This cross-sectional study was conducted at the Mulago Assessment Centre in Mulago National Referral Hospital, Kampala, Uganda, the country’s main tertiary referral and teaching hospital serving patients from Kampala, surrounding districts, and neighbouring countries. The centre receives about 150 outpatients daily. The study enrolled individuals presenting with gastrointestinal symptoms suggestive of ulcer disease—such as abdominal pain, bloating, nausea, or vomiting—who sought care during the study period (1^st^ October to 30^th^ November 2024). Data were collected using a semi-structured questionnaire capturing demographic, lifestyle, and clinical information, alongside stool sample analysis for *H. pylori* detection.

### Sampling procedure and sample size determination

Participants were selected using consecutive sampling, enrolling all eligible patients who presented to the Mulago Assessment Centre with gastrointestinal symptoms and were clinically diagnosed with *H. pylori* infection or gastrointestinal ulcers. This approach minimized selection bias by including patients in the order they appeared, ensuring a representative sample of symptomatic cases. Enrolled participants provided stool samples for laboratory confirmation of *H. pylori* infection and completed a questionnaire capturing demographic characteristics, lifestyle factors, medical history, and gastrointestinal symptoms. The required sample size was determined using the formula for cross-sectional studies by Kish–Leslie, assuming a 95% confidence level, 5% margin of error, and a prevalence of infection of 29.9% based on a previous study in rural Uganda (12). This yielded a minimum sample size of 322 participants, which, after adjusting for a 10% non-response rate, resulted in a final target sample size of 353 participants.

Eligible participants were patients aged 16 years and above presenting with symptoms of gastrointestinal ulcer disease such as abdominal pain, dyspepsia, or gastrointestinal bleeding and provided consent. Patients were excluded if they were critically ill, had taken antibiotics in the past month, or had a history of gastric surgery, gastric malignancy, or prior cancer treatment. Those with severe comorbidities or gastrointestinal conditions likely to confound results were also excluded.

### Sample and data collection procedure

Stool samples were collected from each participant using sterile, labeled containers with buffer solution, following standardized instructions to obtain a small portion (approximately 50–100 mg) of fresh stool. Participants were guided on proper collection to minimize contamination and delivered the samples to the laboratory for analysis. Helicobacter pylori antigen detection was performed using the SD H. pylori Stool Antigen ELISA cassette (Standard Diagnostics Inc., South Korea), with results interpreted according to manufacturer instructions: two red bands indicated a positive result, one red band at the control line indicated a negative result, and absence of a control line indicated an invalid test (13). Laboratory procedures were conducted by trained personnel under the supervision of the principal investigator to ensure accuracy, with repeat testing performed when necessary. Concurrently, a semi-structured questionnaire was administered by trained research assistants to collect demographic data, lifestyle factors, medical history, and gastrointestinal symptoms, which were subsequently managed, coded, and analyzed using Excel and STATA Version 15.0.

### Data management and analysis

The questionnaire was pretested and revised, then administered by two trained research assistants with laboratory backgrounds. The assistants received two days of training on data collection using the Kobo Collect tool, operation of the online questionnaire designed for the study, and sample preparation procedures, to ensure data accuracy and participant confidentiality. Completed forms were checked daily, entered into Excel for cleaning and coding, securely stored, and exported to STATA Version 15.0 for analysis, with backups maintained. Laboratory quality was ensured through control samples, standardized procedures, and proper stool sample handling to preserve integrity and reliability of *H. pylori* detection.

Data analysis was conducted at univariable, bivariable, and multivariable levels. Descriptive statistics summarized participant characteristics using means, standard deviations, frequencies, percentages, and proportions, presented in tables and charts. The prevalence of *H. pylori* infection was calculated as the proportion of participants testing positive. Bivariate analysis identified potential risk factors (p□<□0.2) for inclusion in the multivariable model, alongside variables recognized in the literature. Multicollinearity was assessed using variance inflation factors, and a modified Poisson regression model was employed to estimate prevalence rate ratios with 95% confidence intervals, given the relatively high outcome prevalence. A stepwise forward and backward approach was used to build the model, adding or removing variables based on significance and model fit to identify independent predictors of H. pylori infection.

### Ethical considerations

Ethical approval was obtained from the Makerere University School of Public Health Higher Degrees Committee. Written informed consent was obtained from all participants prior to data and stool sample collection. Privacy, confidentiality, and voluntary participation were ensured, with benefits and risks explained. The study adhered to ethical principles of beneficence, justice, and scientific integrity, and no harm was anticipated beyond the time required for participation.

## Results

### Socio-demographic characteristics of the study participants

The majority of participants were youth aged 16-35 years 242 (69%), females 204 (58%) and had attained secondary education 123 (35%). Most of the participants resided in peri-urban areas 262 (74%), were single 194 (55%), and unemployed 291 (82%). The majority of households had incomes of less than 139 USD (at a rate of 3,600 UGX) 268 (76%), with mainly 1 to 5 household members (Table 1).

**Table 1:**
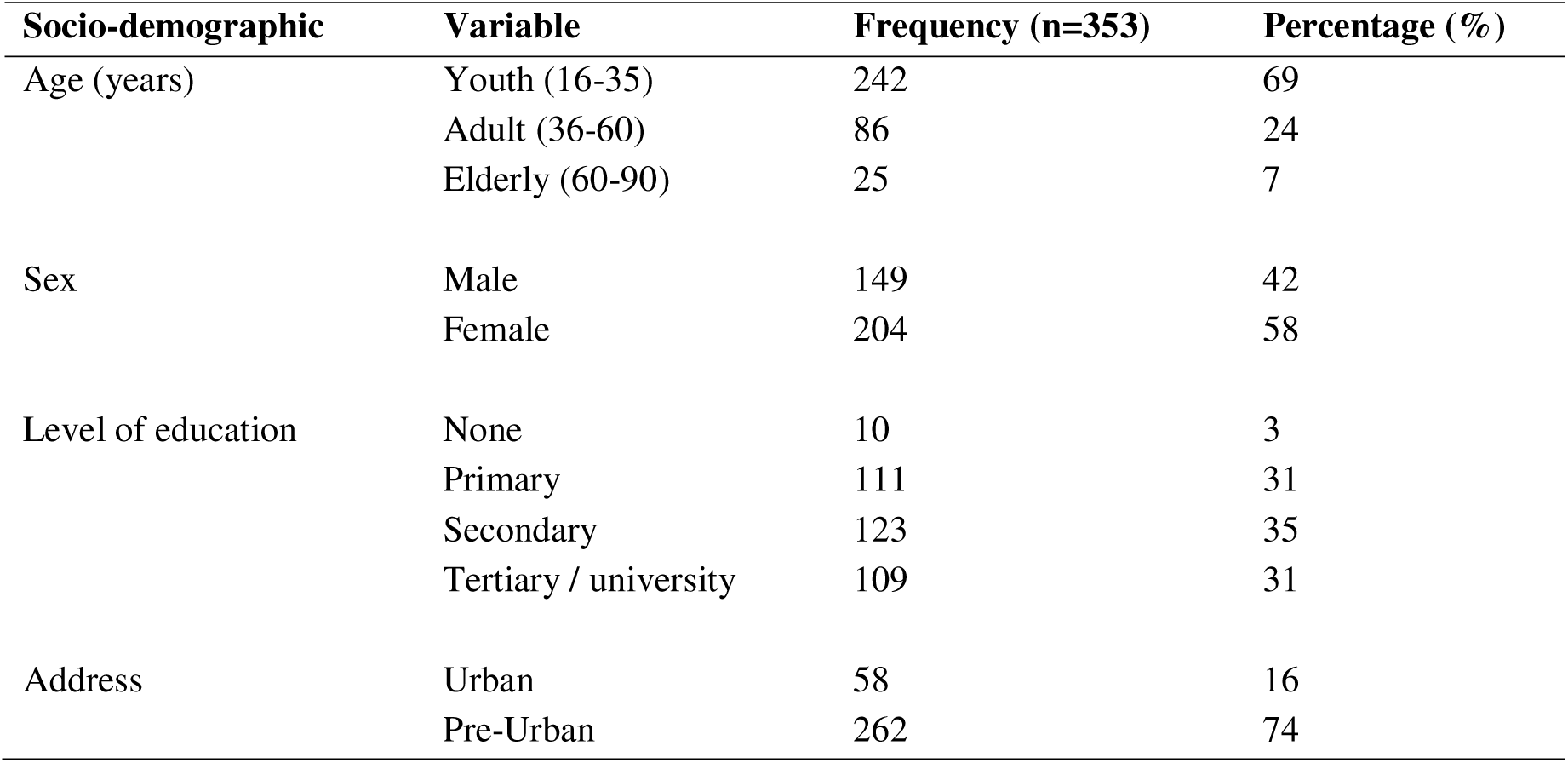
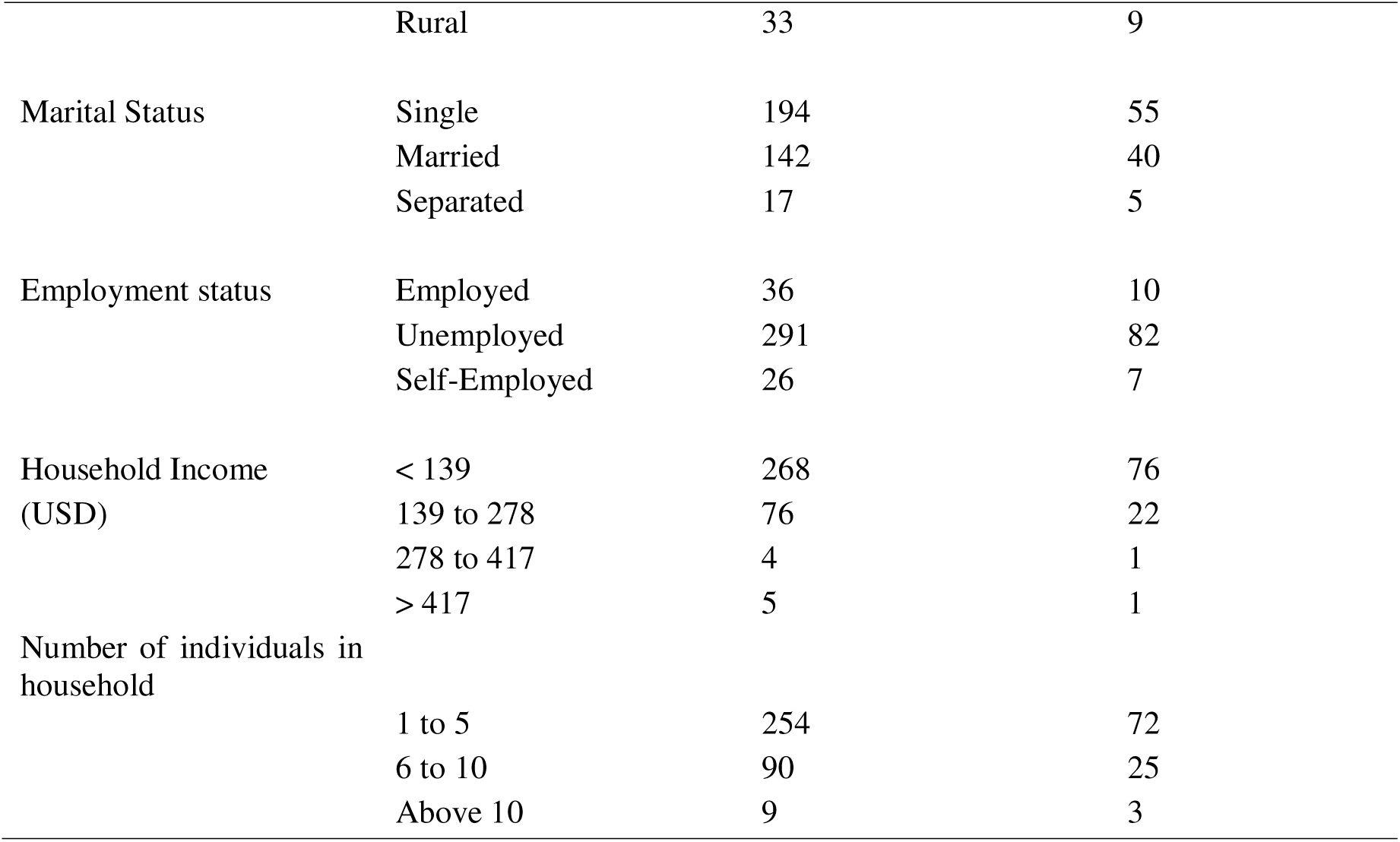
Socio Demographic characteristics of study participants.

### Lifestyle and Dietary Habits of the Participants

The study revealed that almost all (99.4%) participants were non-smokers, did not consume alcohol (87%), but consumed spicy foods weekly (45%). Most participants (80.5%) always practiced hand washing, sourced food from more than one place (75.7%), and most obtained water from more than one source (81%) (Table 2).

**Table 2:**
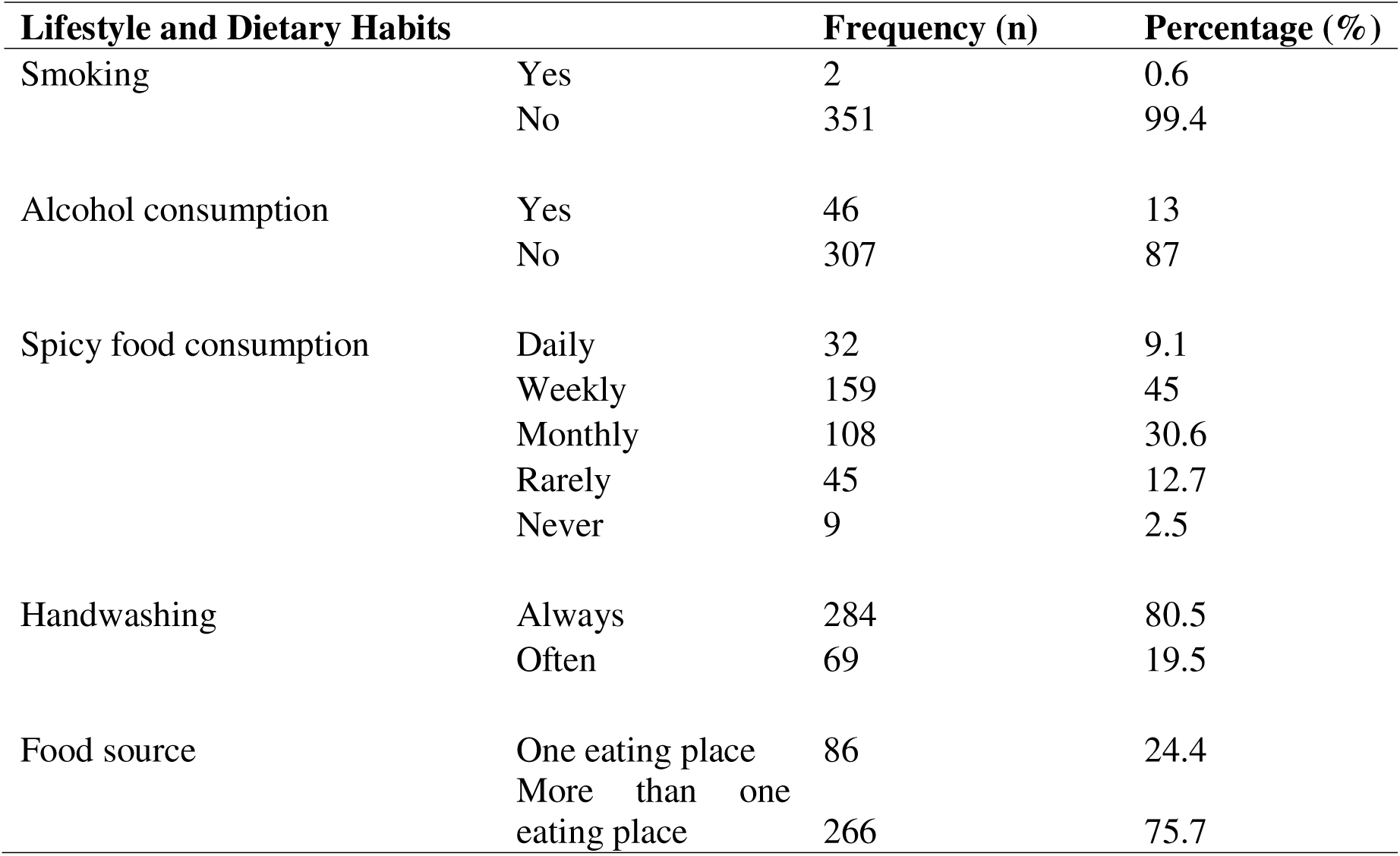
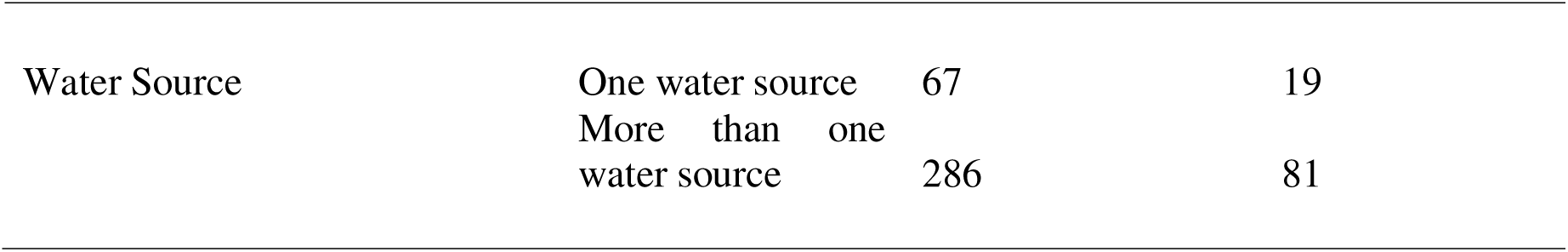
Lifestyle and Dietary Habits of the Participants.

| Lifestyle and Dietary Habits |  | Frequency (n) | Percentage (%) |
| --- | --- | --- | --- |
| Smoking | Yes | 2 | 0.6 |
|  | No | 351 | 99.4 |
| Alcohol consumption | Yes | 46 | 13 |
|  | No | 307 | 87 |
| Spicy food consumption | Daily | 32 | 9.1 |
|  | Weekly | 159 | 45 |
|  | Monthly | 108 | 30.6 |
|  | Rarely | 45 | 12.7 |
|  | Never | 9 | 2.5 |
| Handwashing | Always | 284 | 80.5 |
|  | Often | 69 | 19.5 |
| Food source | One eating place | 86 | 24.4 |
|  | More than one eating place | 266 | 75.7 |
| Water Source | One water source | 67 | 19 |
|  | More than one water source | 286 | 81 |

### Medical History of the participants

The majority (54.4%) of participants had never been diagnosed with a gastrointestinal tract (GIT) ulcer, 95.2% had been tested for *H. pylori* infection, and 87.3% reported having received treatment for it. Regarding general health and medications, only 6.8% were currently taking any medications. Most participants (89.8%) had used non-steroidal anti-inflammatory drugs (NSAIDs) such as aspirin or ibuprofen in the previous year, while only 18.7% reported having a family member diagnosed with *H. pylori* infection or GIT ulcers (Table 3).

**Table 3:**
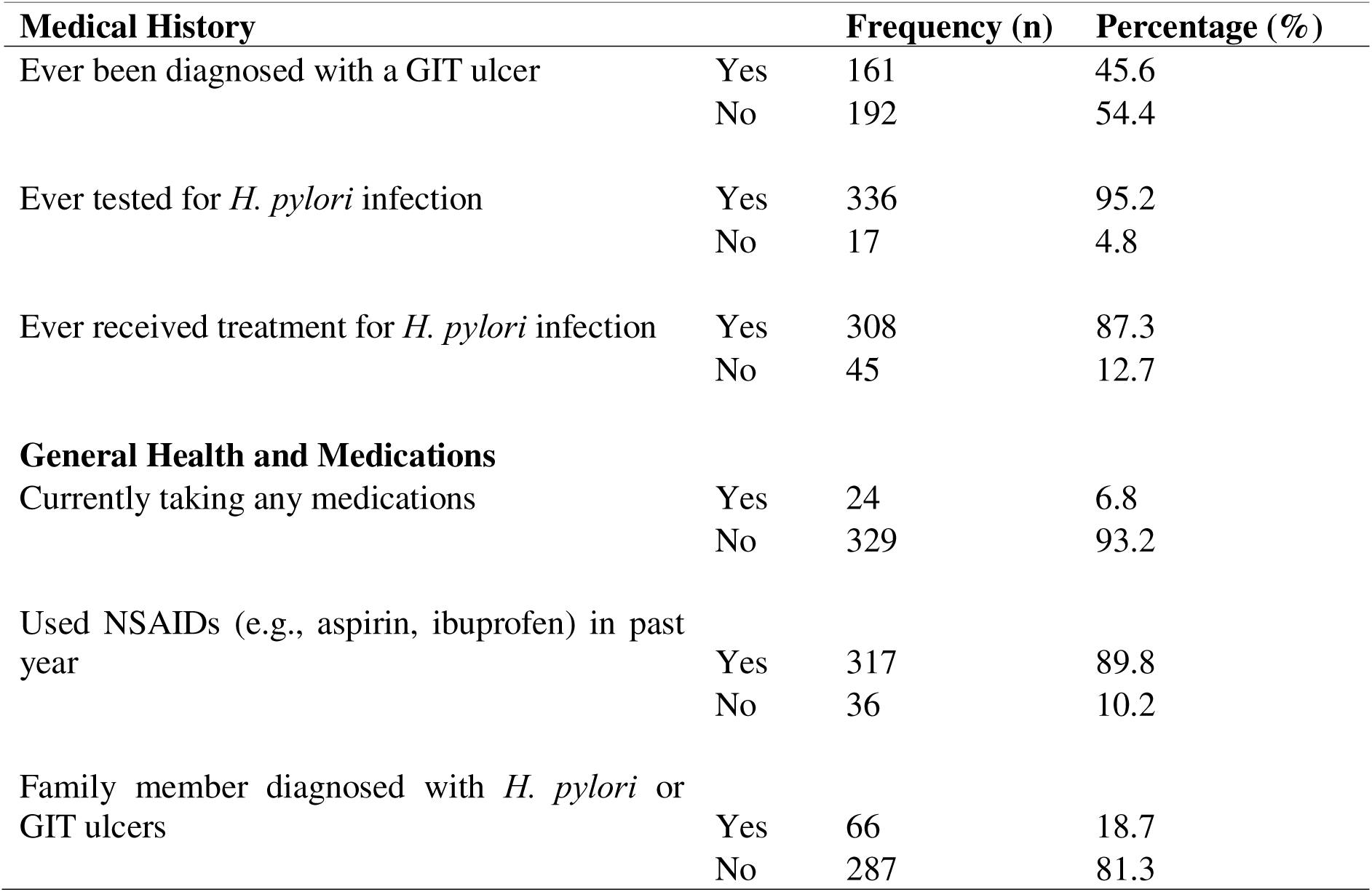
Medical History of the Participants.

| Medical History |  | Frequency (n) | Percentage (%) |
| --- | --- | --- | --- |
| Ever been diagnosed with a GIT ulcer | Yes | 161 | 45.6 |
|  | No | 192 | 54.4 |
| Ever tested for *H. pylori* infection | Yes | 336 | 95.2 |
|  | No | 17 | 4.8 |
| Ever received treatment for *H. pylori* infection | Yes | 308 | 87.3 |
|  | No | 45 | 12.7 |
| **General Health and Medications** |  |  |  |
| Currently taking any medications | Yes | 24 | 6.8 |
|  | No | 329 | 93.2 |
| Used NSAIDs (e.g., aspirin, ibuprofen) in past year | Yes | 317 | 89.8 |
|  | No | 36 | 10.2 |
| Family member diagnosed with *H. pylori* or GIT ulcers | Yes | 66 | 18.7 |
|  | No | 287 | 81.3 |

### Prevalence of *H. pylori* among patients with gastrointestinal symptoms at Mulago Hospital

Out of the 353 participants tested, 308 were positive for *H. pylori* hence a prevalence of 87.3%.

### Bivariate analysis of Pre-disposing factors for *H. pylori* among patients with gastrointestinal symptoms at Mulago Hospital

#### Demographic factors associated with *H. pylori*

Among the demographic factors assessed, employment and self-employment were statistically significant (cPRR=0.793, 95% CI: 0.645-0.974, p=0.027) and (cPRR=0.717, 95% CI: 0.541-0.952, p=0.021) respectively. Additionally, respondents with more than five financial dependents showed statistical significance (cPRR=1.109, 95% CI: 1.032- 1.193, p=0.005) and hence were considered for multivariable analysis (Table 4).

**Table 4:**
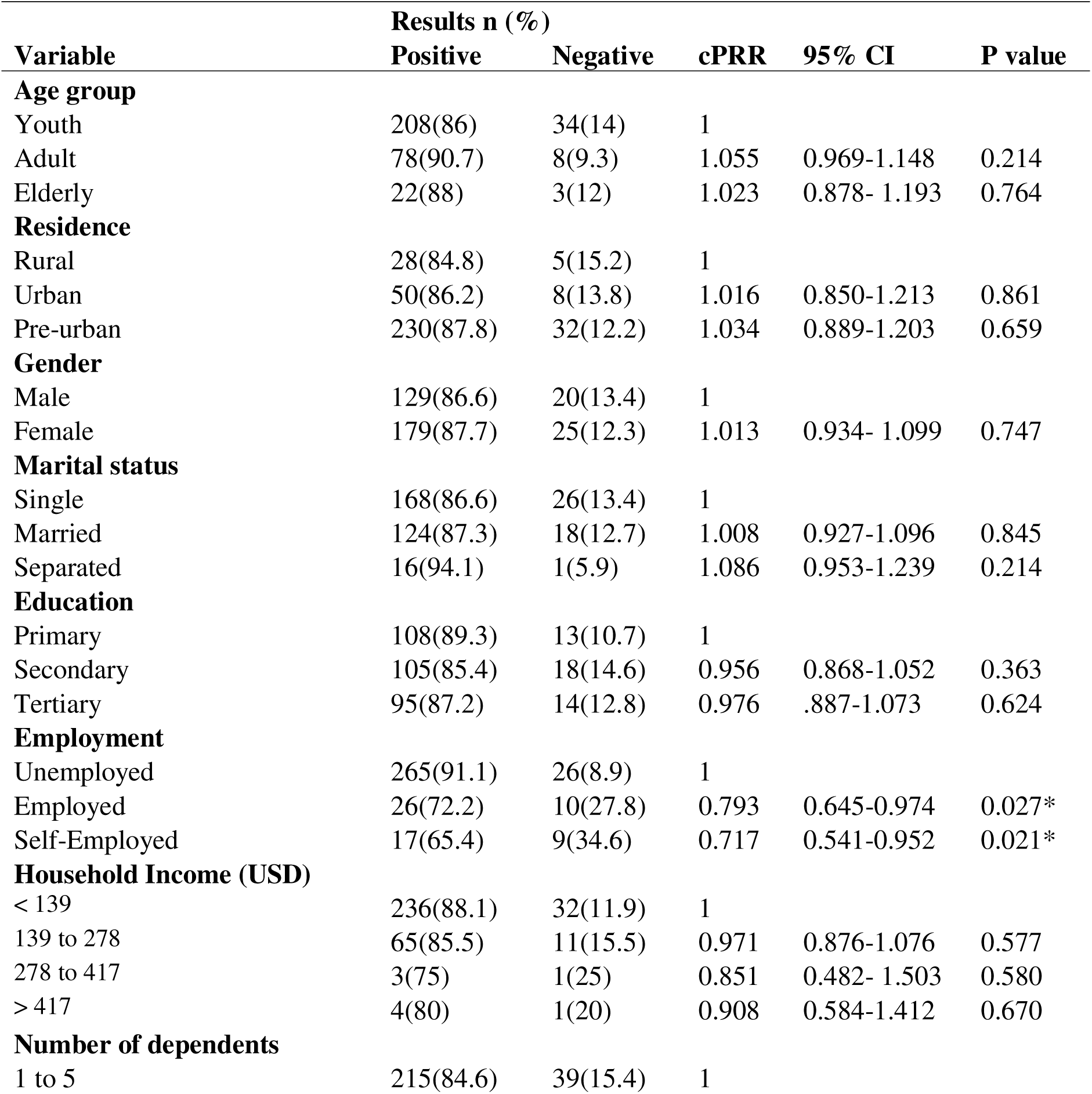
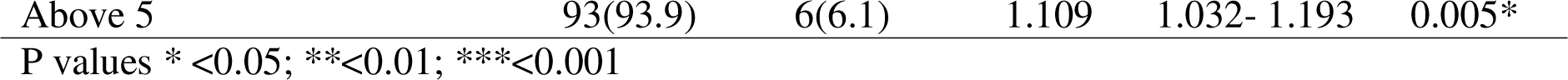
Demographic factors associated with H. pylori.

#### Lifestyle and Dietary Habits factors associated with *H. pylori*

At bivariate analysis, none of the lifestyle and dietary habits were statistically significant (p<0.2) hence they were not considered for multivariable analysis (Table 5).

**Table 5:** Lifestyle and Dietary Habits Factors Associated with H. pylori.

| Lifestyle and Dietary Habits | Results n (%) |  | cPRR | 95% CI | P value |
| --- | --- | --- | --- | --- | --- |
|  | Positive | Negative |  |  |  |
| **Smoking** |  |  |  |  |  |
| Yes | 1(50) | 1(50) | 1 |  |  |
| No | 307(87.5) | 44(12.5) | 1.749 | 0.436- 7.011 | 0.430 |
| **Alcohol consumption** |  |  |  |  |  |
| Yes | 38(82.6) | 8(17.4) | 1 |  |  |
| No | 270(87.9) | 37(12.1) | 1.064 | 0.926-1.223 | 0.378 |
| **Spicy food consumption** |  |  |  |  |  |
| Daily | 28(87.5) | 4(12.5) | 1 |  |  |
| Weekly | 141(88.7) | 18(11.3) | 1.013 | 0.878- 1.168 | 0.854 |
| Monthly | 91(84.3) | 17(15.7) | 0.962 | 0.825-1.123 | 0.632 |
| Rarely | 48(88.9) | 6(11.1) | 1.015 | 0.864- 1.194 | 0.849 |
| **Handwashing** |  |  |  |  |  |
| Always | 247(87) | 37(13) | 1 |  |  |
| Often | 61(88.4) | 8(11.6) | 1.016 | 0.922-1.119 | 0.74 |
| **Food source** |  |  |  |  |  |
| One eating place | 77(89.5) | 9(10.5) | 1 |  |  |
| More than one eating place | 231(86.5) | 36(13.5) | 0.966 | 0.886- 1.053 | 0.437 |
| **Water Source** |  |  |  |  |  |
| One water source | 61(91) | 6(9) | 1 |  |  |
| More than one water source | 247(86.4) | 39(13.6) | 0.948 | 0.868-1.036 | 0.241 |

P values \* <0.05; \*\*<0.01; \*\*\*<0.001

#### Medical history factors associated with *H. pylori*

Regarding medical history-related factors, ever been diagnosed with a GIT ulcer (cPRR=1.074, 95% CI: 0.993-1.162, p=0.072), having ever tested for *H. pylori* infection (cPRR=1.366, 95% CI: 0.959-1.945, p=0.084), ever received treatment for *H. pylori* infection (cPRR=3.603, 95% CI: 2.217-5.857, p<0.001), having used NSAIDS (e.g., aspirin, ibuprofen) in past year (cPRR=1.285, 95% CI: 1.031-1.602, p=0.025), and having a family member diagnosed with *H. pylori* or GIT ulcers (cPRR=1.14, 95% CI: 1.069-1.216, p<0.001) were considered for multivariate analysis (Table 6).

**Table 6:**
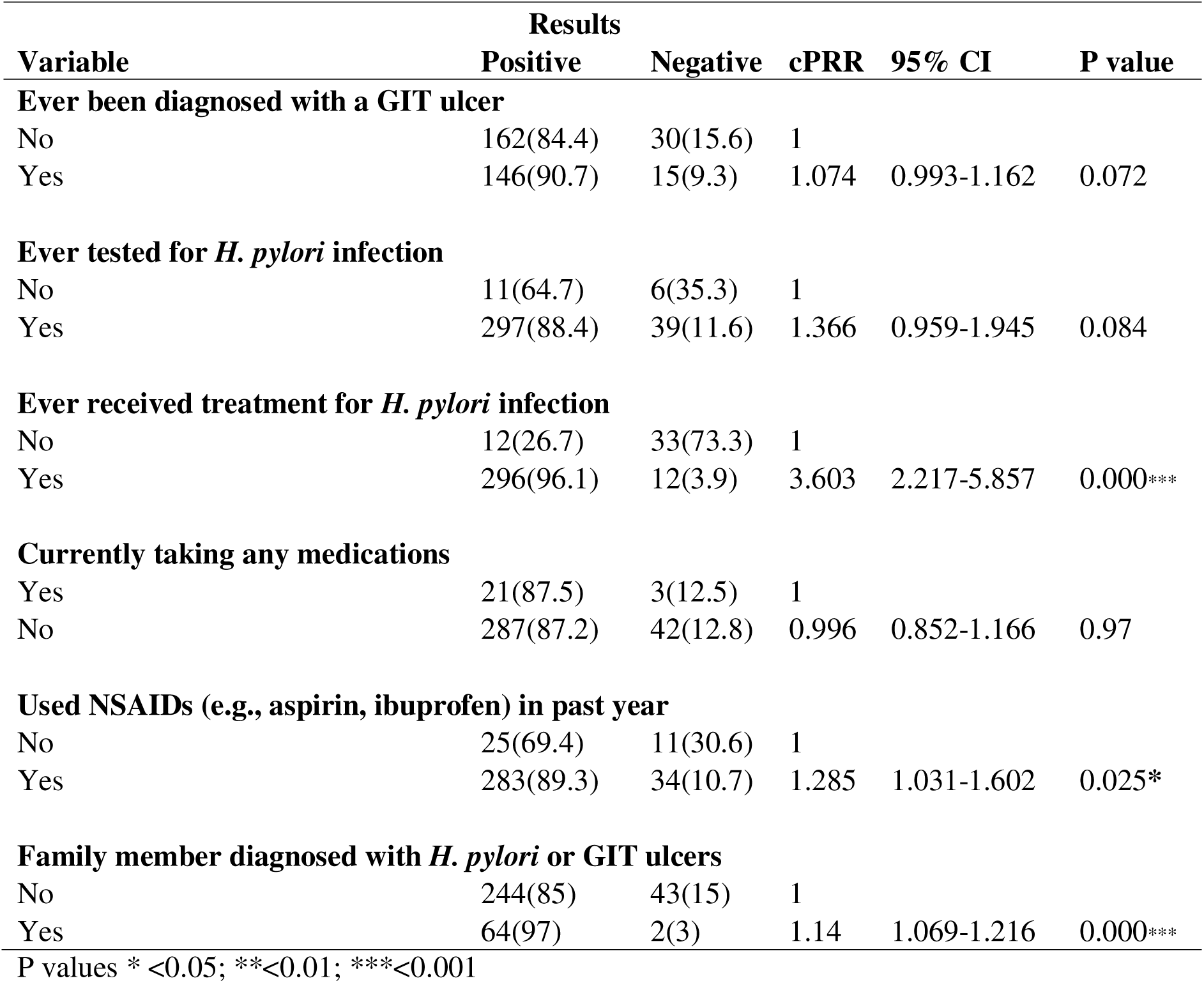
Bivariate analysis of medical history factors associated with H. pylori.

| Variable | Results |  | cPRR | 95% CI | P value |
| --- | --- | --- | --- | --- | --- |
|  | Positive | Negative |  |  |  |
| **Ever been diagnosed with a GIT ulcer** |  |  |  |  |  |
| No | 162(84.4) | 30(15.6) | 1 |  |  |
| Yes | 146(90.7) | 15(9.3) | 1.074 | 0.993-1.162 | 0.072 |
| **Ever tested for *H. pylori* infection** |  |  |  |  |  |
| No | 11(64.7) | 6(35.3) | 1 |  |  |
| Yes | 297(88.4) | 39(11.6) | 1.366 | 0.959-1.945 | 0.084 |
| **Ever received treatment for *H. pylori* infection** |  |  |  |  |  |
| No | 12(26.7) | 33(73.3) | 1 |  |  |
| Yes | 296(96.1) | 12(3.9) | 3.603 | 2.217-5.857 | 0.000*** |
| **Currently taking any medications** |  |  |  |  |  |
| Yes | 21(87.5) | 3(12.5) | 1 |  |  |
| No | 287(87.2) | 42(12.8) | 0.996 | 0.852-1.166 | 0.97 |
| **Used NSAIDs (e.g., aspirin, ibuprofen) in past year** |  |  |  |  |  |
| No | 25(69.4) | 11(30.6) | 1 |  |  |
| Yes | 283(89.3) | 34(10.7) | 1.285 | 1.031-1.602 | 0.025* |
| **Family member diagnosed with *H. pylori* or GIT ulcers** |  |  |  |  |  |
| No | 244(85) | 43(15) | 1 |  |  |
| Yes | 64(97) | 2(3) | 1.14 | 1.069-1.216 | 0.000*** |

P values \* <0.05; \*\*<0.01; \*\*\*<0.001

### Multivariate analysis of factors associated with *H. Pylori* infection among patients with gastro-intestinal symptoms attending Mulago Hospital

After controlling for confounding with age at multivariable analysis, the significant variables found positively, associated with testing positive for *H. pylori*, were participants with more than five financial dependents were more likely to test positive compared to those with one to five dependents (aPRR=1.104, 95% CI: 1.025–1.189, p = 0.008). Those who had ever received treatment for *H. pylori* infection were substantially more likely to test positive compared to those who had not received treatment (aPRR=3.459, 95% CI: 2.138–5.595, p < 0.001). Additionally, participants with a family member diagnosed with *H. pylori* infection or gastrointestinal tract (GIT) ulcers were more likely to test positive compared to those without such a family history (aPRR=1.135, 95% CI: 1.055–1.221, p = 0.001) (Table 7).

**Table 7:**
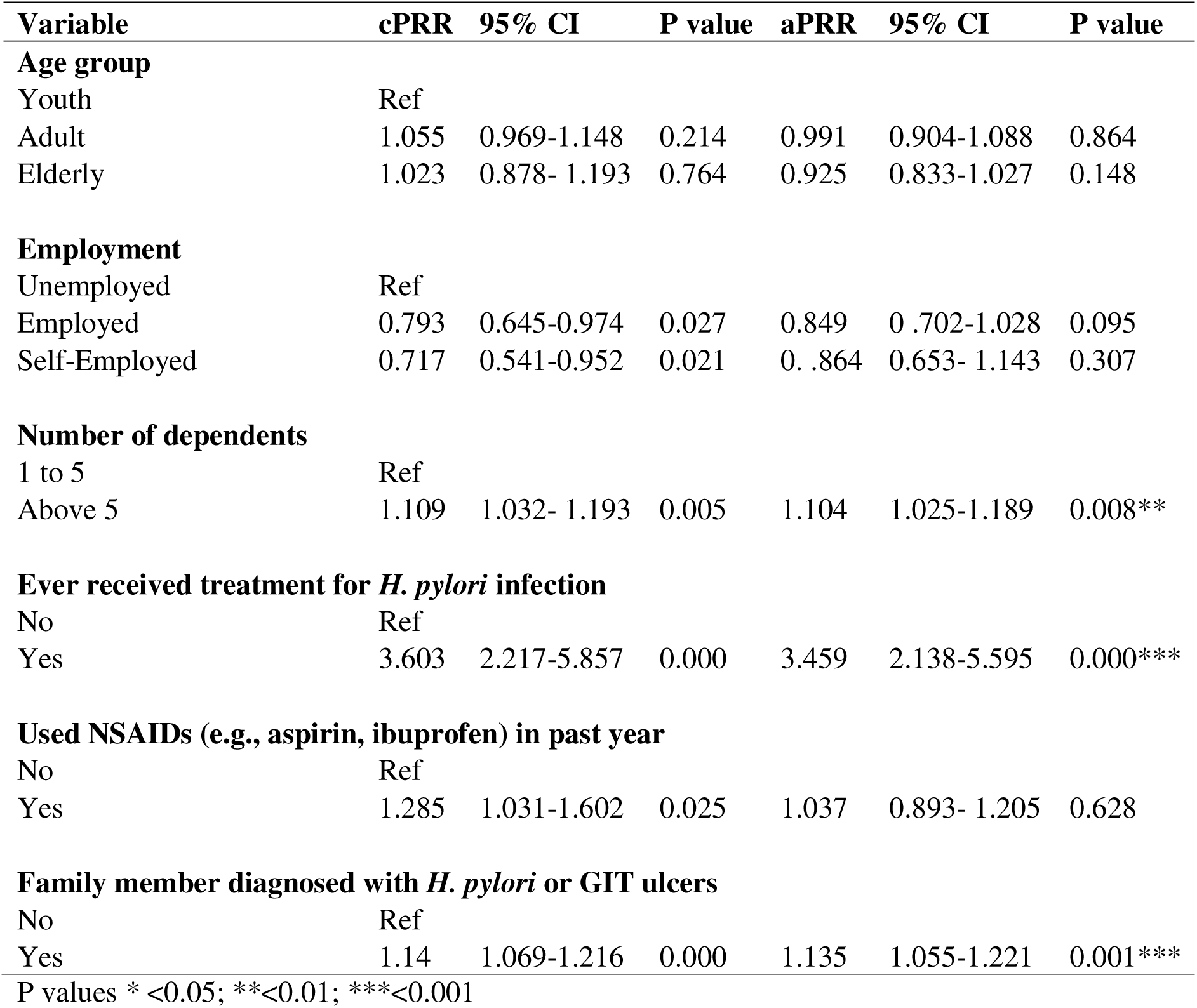
Multivariate analysis of factors associated with H. Pylori infection among patients with gastro-intestinal symptoms attending Mulago Hospital.

| Variable | cPRR | 95% CI | P value | aPRR | 95% CI | P value |
| --- | --- | --- | --- | --- | --- | --- |
| **Age group** |  |  |  |  |  |  |
| Youth | Ref |  |  |  |  |  |
| Adult | 1.055 | 0.969-1.148 | 0.214 | 0.991 | 0.904-1.088 | 0.864 |
| Elderly | 1.023 | 0.878- 1.193 | 0.764 | 0.925 | 0.833-1.027 | 0.148 |
| **Employment** |  |  |  |  |  |  |
| Unemployed | Ref |  |  |  |  |  |
| Employed | 0.793 | 0.645-0.974 | 0.027 | 0.849 | 0.702-1.028 | 0.095 |
| Self-Employed | 0.717 | 0.541-0.952 | 0.021 | 0.864 | 0.653- 1.143 | 0.307 |
| **Number of dependents** |  |  |  |  |  |  |
| 1 to 5 | Ref |  |  |  |  |  |
| Above 5 | 1.109 | 1.032- 1.193 | 0.005 | 1.104 | 1.025-1.189 | 0.008** |
| **Ever received treatment for *H. pylori* infection** |  |  |  |  |  |  |
| No | Ref |  |  |  |  |  |
| Yes | 3.603 | 2.217-5.857 | 0.000 | 3.459 | 2.138-5.595 | 0.000*** |
| **Used NSAIDs (e.g., aspirin, ibuprofen) in past year** |  |  |  |  |  |  |
| No | Ref |  |  |  |  |  |
| Yes | 1.285 | 1.031-1.602 | 0.025 | 1.037 | 0.893- 1.205 | 0.628 |
| **Family member diagnosed with *H. pylori* or GIT ulcers** |  |  |  |  |  |  |
| No | Ref |  |  |  |  |  |
| Yes | 1.14 | 1.069-1.216 | 0.000 | 1.135 | 1.055-1.221 | 0.001*** |

P values \* <0.05; \*\*<0.01; \*\*\*<0.001

## Discussion

This study found an exceptionally high prevalence of *H. pylori* infection (87.3%) among symptomatic patients attending Mulago Hospital, highlighting a substantial and ongoing public health challenge in Uganda. These findings reflect widespread transmission within households, persistent reinfection or treatment failure, and socio-environmental conditions that continue to facilitate the spread of the bacterium. Given that Mulago Hospital serves as Uganda’s national referral centre—receiving complex cases from Kampala, other regions of Uganda, and neighbouring countries—this high prevalence should be interpreted within this specific clinical context and may not be representative of the general population or primary healthcare settings. Overall, the results underscore the need for strengthened public health interventions, improved diagnostic and treatment strategies, and community-level measures to reduce the burden of *H. pylori* and its long-term complications.

The prevalence reported in this study is considerably higher than that observed in previous Ugandan studies, likely reflecting differences in study populations, settings, and diagnostic methods. Namyalo et al. reported a prevalence of 35.7% among private clinic attendees (11), while Baingana et al. (2014) found 45.2% among pregnant women (14). Diagnostic variation plays a critical role, as Kakooza (2021) reported a lower prevalence of 29.2% using antibody tests that indicate past exposure rather than active infection, unlike the stool antigen testing employed in this study (15). Similarly, the lower prevalence reported among children in Mbarara by Aitila et al. (2019) likely reflects shorter exposure duration (16). These contextual and methodological differences underscore the substantial burden of *H. pylori* infection among symptomatic adults seeking care in urban tertiary referral facilities. While this study provides important evidence on the burden of *H. pylori* among patients presenting with gastrointestinal symptoms at a tertiary-care hospital in Uganda, further research is needed to establish prevalence in primary healthcare settings and the general population. The findings reaffirm *H. pylori* infection as a significant public health and clinical concern, particularly in referral hospitals where patients often present with more advanced or persistent disease, and highlight the need for broader population-based studies to inform national screening, management, and prevention guidelines. Global evidence (1) and local studies (17, 18) further emphasize the importance of targeted prevention, surveillance, and evidence-based treatment protocols tailored to Uganda’s antibiotic resistance patterns.

Participants with more than five income dependents were more likely to test positive, due to overcrowding, shared living spaces, and limited resources that facilitate person-to-person transmission and shared living conditions that facilitate transmission through oral-oral or fecal-oral routes. This aligns with findings from Uganda and other countries, including observations from a study in South Western Uganda that found *H. pylori* infections were more common among children in crowded households (16). Similar trends have been observed in other countries, including China (19), Turkey (20), and Germany (21) where familial exposure is considered a major source of transmission especially between parents and children, These findings highlight the need for targeted interventions aimed at improving hygiene practices in larger households, alongside educational programs that emphasize proper sanitation, safe food handling, and strategies to reduce transmission in crowded living conditions. Strengthening community- and household-level interventions could significantly reduce the spread of *H. pylori* and its associated health risks, particularly in densely populated settings where close contact and shared facilities increase the likelihood of transmission.

Participants previously treated for *H. pylori* were more likely to test positive again, suggesting reinfection, recurrence, or incomplete eradication. These challenges may stem from persistent exposure, poor treatment adherence, or antibiotic resistance. Similar trends have been reported in studies assessing treatment outcomes (22, 23). Family-based management strategies—such as screening and treating household members, promoting good hygiene practices, improving access to clean water and sanitation, and ensuring appropriate follow-up care—have the potential to reduce reinfection rates by addressing shared household risk factors. These findings underscore the importance of comprehensive interventions that integrate patient education, tailored treatment regimens, improved adherence, monitoring of antibiotic resistance, and screening of close contacts. Such coordinated patient-, household-, and community-level approaches are essential to interrupt ongoing transmission and break the cycle of reinfection.

Participants with a family history of *H. pylori* infection or gastrointestinal ulcers were more likely to test positive, further supporting evidence that transmission is often intra-household. Close contact, shared utensils, and common sanitation challenges contribute to this pattern, as described by Borka Balas et al. (2022) (24). Screening and treating family members of infected individuals could disrupt transmission pathways and reduce household reinfection risks. Family-based management strategies, which target all infected or at-risk members within a household, offer a more comprehensive approach to minimizing infection rates. Public health efforts should prioritize such strategies, particularly in high-prevalence regions, as they not only address individual cases but also help prevent broader community transmission and the long-term health impacts of *H. pylori*.

### Strengths and limitations of the study

The study used consecutive sampling, which may have introduced selection bias and limited the generalizability of the findings, as only patients available during the data collection period were included. Conducting the study at Mulago Hospital, a national referral centre, also meant that participants were more likely to present with severe gastrointestinal symptoms, contributing to the high *H. pylori* prevalence and limiting applicability to community or primary care settings. Self-reported information on household conditions, diet, and treatment history may have been affected by recall bias, and the lack of qualitative data prevented deeper insight into transmission factors. Despite these limitations, the study’s major strength is its implementation at a large referral centre serving a diverse population, providing important epidemiological evidence to guide clinical practice and inform health policy.

## Conclusion

The study found a very high *H. pylori* prevalence (9 in 10 patients) with gastrointestinal symptoms at Mulago National Referral Hospital. Infection was strongly associated with having more than five income dependents, previous *H. pylori* treatment, and a family history of infection or gastrointestinal ulcers, underscoring the role of household and individual factors in transmission. Addressing these issues through better housing, awareness of familial risk, standardized treatment, and follow-up is crucial to reducing the infection burden of *H. Pylori.* These findings highlight *H. pylori* as a major clinical and public health concern in urban, resource-limited settings. Further research in primary healthcare and community populations is needed to guide wider prevention and management efforts.

## Data Availability

All data produced in the present study are available upon reasonable request to the authors

## Acknowledgement

I would like to express my heartfelt gratitude to my husband and family for their unwavering support, encouragement, and understanding throughout this journey. I am also profoundly thankful to my colleagues in the Master of Public Health Distance Education program and our discussion group, whose collaboration, constructive feedback, and shared knowledge have been invaluable in shaping this work. Finally, I extend my sincere appreciation to all those who contributed, directly (Dr. Elizabeth Nabiwemba) or indirectly, to the successful completion of this study.

---

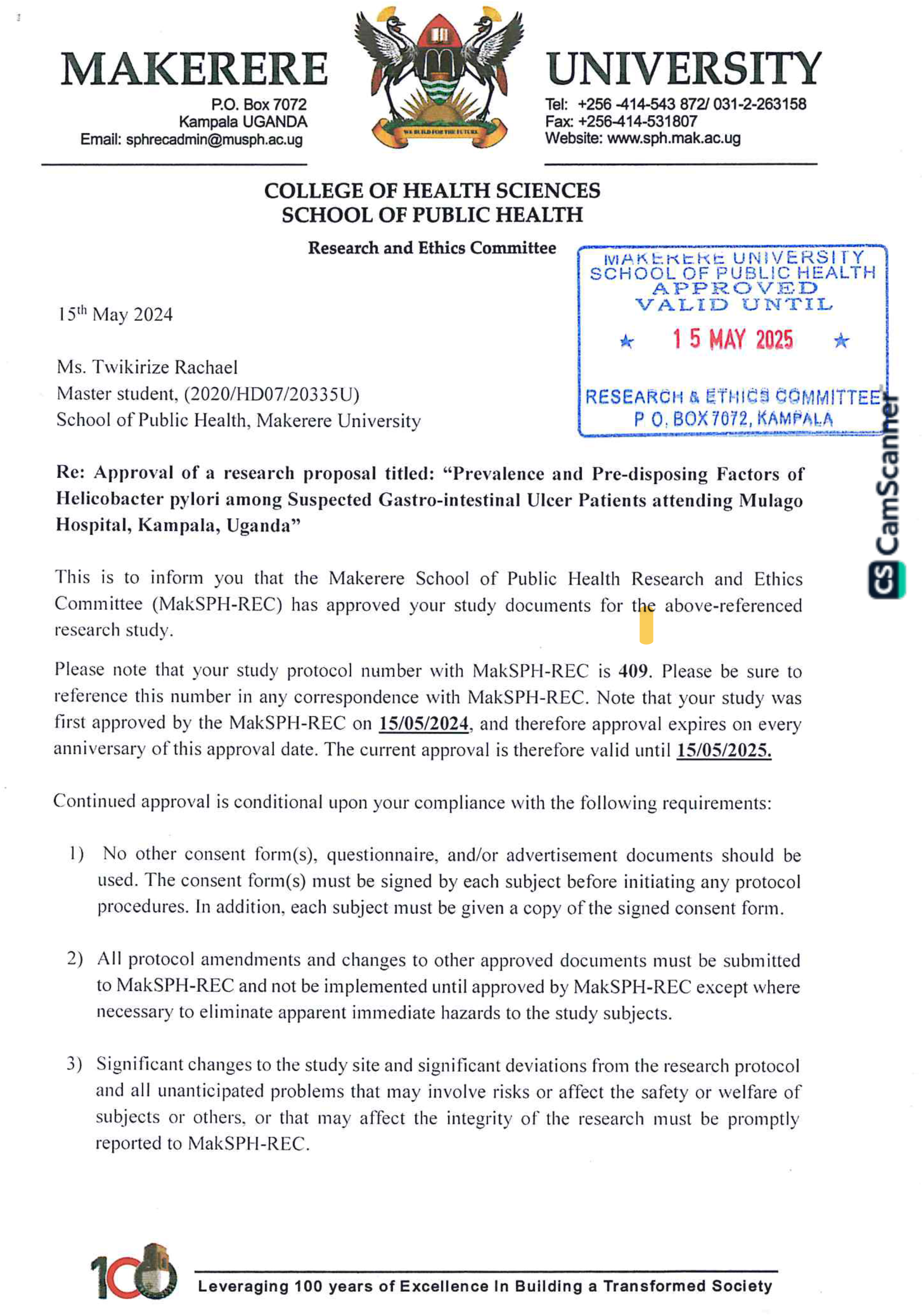
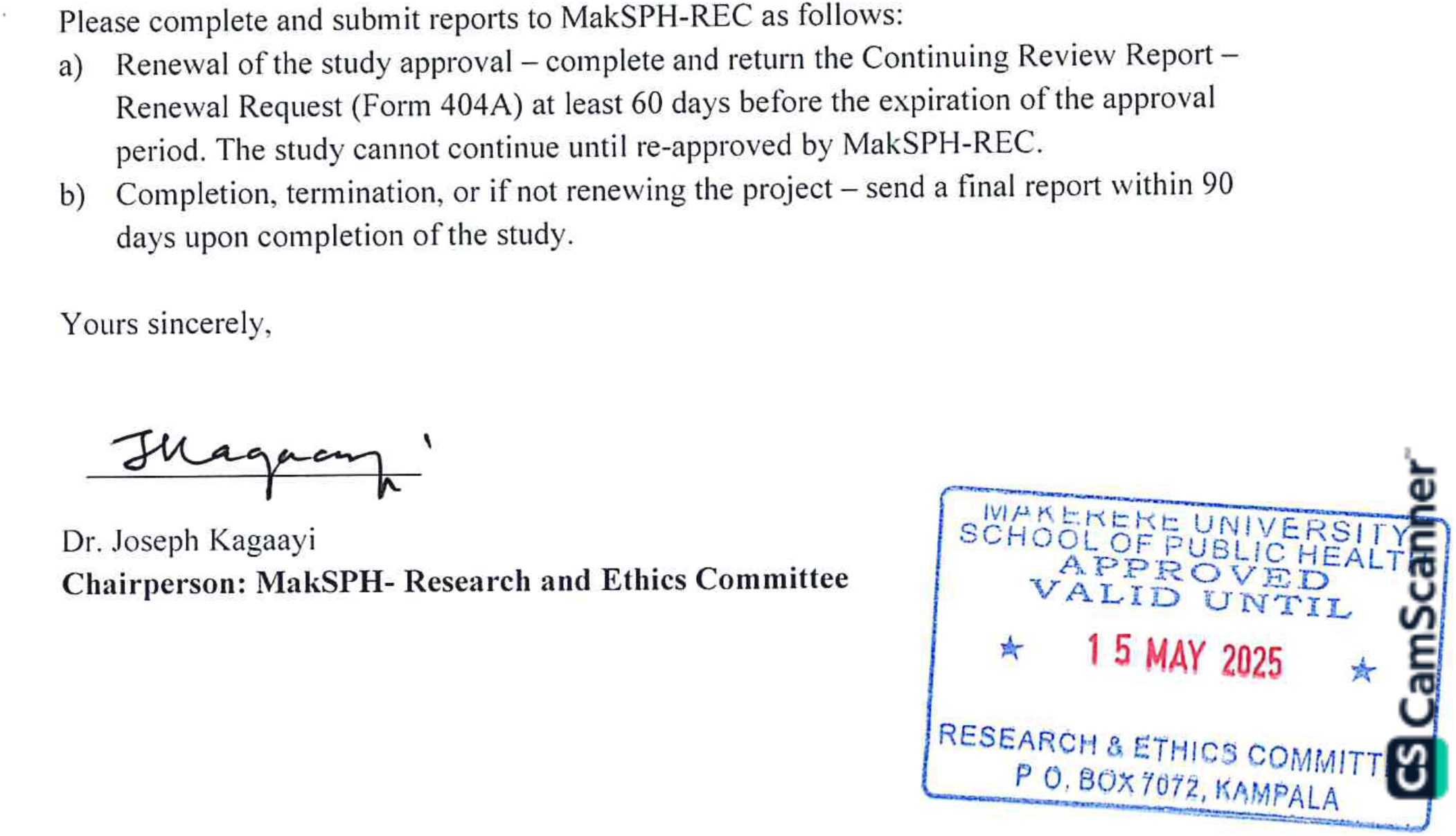

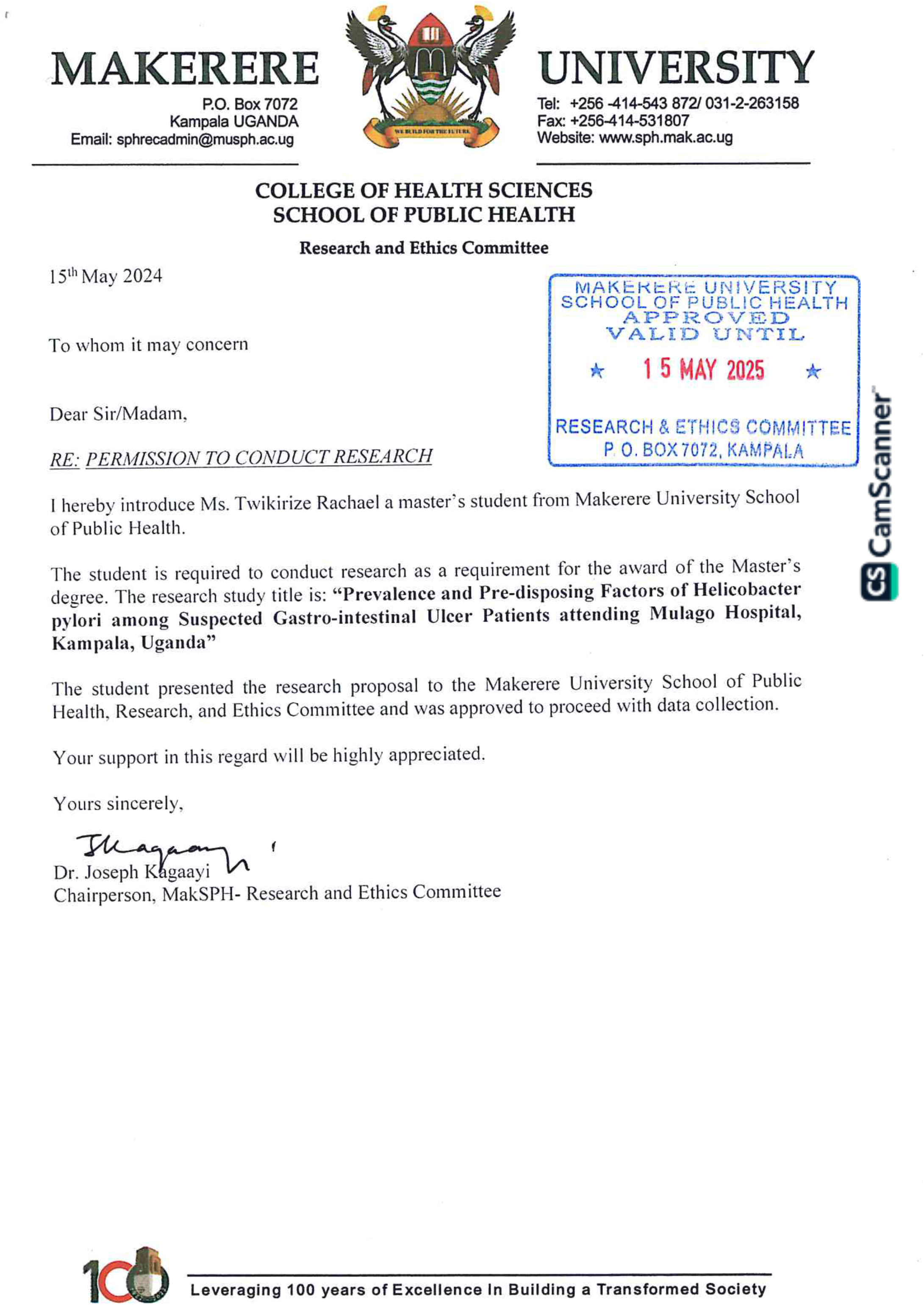
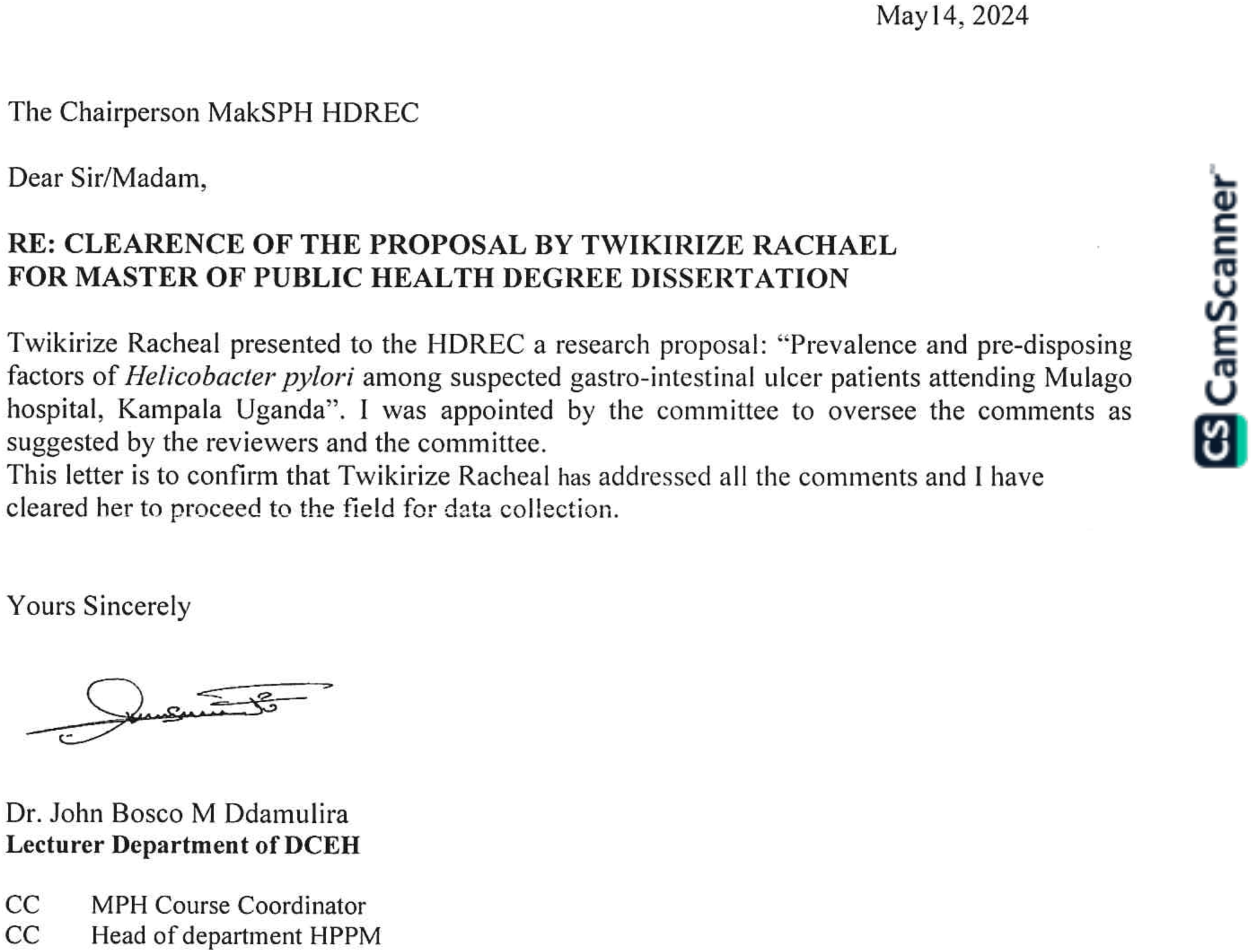

## Notes

### Competing Interest Statement

The authors have declared no competing interest.

### Funding Statement

This study did not receive any funding

### Author Declarations

Ethical approval was obtained from the Makerere University School of Public Health Higher Degrees Research and Ethics Committee

